# The Role of Community Health Workers in Oral Health Promotion and the Impact of their services in Sub-Saharan Africa: A Systematic Review

**DOI:** 10.1101/2021.04.06.21255025

**Authors:** Mohammed Azhar Khan, Bernard Ojiambo Okeah, Etheldreda Leinyuy Mbivnjo, Ephraim Kisangala, Aaron Wyn Pritchard

## Abstract

Oral ailments are largely preventable but remain a significant public health concern afflicting nearly half the global population. These conditions account for 220 years of life lost per 100,000 people and about US$500 billion in health-related expenditures. Sub-Saharan Africa bears a significant burden of oral health problems thus exerting additional pressure on the scarce human resources for health. Community healthcare workers (CHWs) could be potentially utilised to bridge the shortage of oral health professionals in sub-Saharan Africa, hence, this systematic review that seeks to explore their current roles in oral health and potential impact on general physical health. This review follows the PRISMA guidelines and databases searched include PubMed, Web of Science, Medline, and CINAHL published between January 2010 and December 2019. Nine studies met the study eligibility criteria. This review established that CHWs perform variable roles cutting across primary, secondary, and tertiary prevention including providing oral hygiene education, recognising common pathologies, and treating oral lesions, administration of tooth extractions, dental pain management, and referral for advanced care. Although this could potentially improve oral health, our review did not establish the extent of the specific impact on general physical health of patients and the burden of oral condition.

## Introduction

“Oral health is multifaceted and includes the ability to speak, smile, smell, taste, touch, chew, swallow, and convey a range of emotions through facial expressions with confidence and without pain, convey a range of emotions through facial expressions with confidence and without pain, discomfort, and disease of the craniofacial complex” ^1^. Oral health is an important aspect of an individual’s life, influencing general health and well-being, psychological ^2^, physiological and social functioning ^1^. Oral health is dynamic (subject to changes in an individual’s expectations, perceptions, experiences and adaptability to conditions) and occurs along a continuum that is subject to the attitudes and values of individuals and communities ^1^.

Many diseases associated with poor oral health, like tooth decay (caries), are largely preventable; yet remain highly prevalent conditions affecting about half of the global human population ^3^. Oral diseases are also responsible for the loss of more than 220 healthy life years per 100,000 people ^4^. In addition, it is estimated that more than US $500 billion was spent globally on managing oral diseases in 2015 ^5^. This amount of money represents a significant strain on the rural populations living where the greatest burden of oral diseases exists but limited access to resources ^3,6^. In rural Sub Saharan Africa (SSA) the population experiences significant barriers that threaten the promotion of oral health ^7,8^ scarcity of oral health care professionals ^7^, long distances to and high costs of accessing oral health services ^7^, ignorance ^9^ as well as unhealthy cultural practices and beliefs ^10^.

There is therefore a need for community-led public health intervention such as the use of community health workers (CHWs) that utilises a preventative approach to tackle the problem. Some researchers have also advocated for the training of CHWs to increase the coverage of oral health programs and address the shortage of trained oral health professionals available to work at the community level ^11^. The implementation of CHWs program can provide a triple benefit to the society including prevention of diseases, promotion of good health practices and basic curative services to the members of the community ^12^. The prospect of early identification and referral of HIV-related oral lesions in CHW led programmes could be key to determining antiretroviral treatment failure ^13^. The new framework for oral health definition also emphasizes access to care as a determining factor and highlights the moderating effect of cultural factors on an individual’s self-evaluation of their oral health ^1^. Moreover, CHWs have been shown to improve the provision of culturally competent healthcare services ^14^ and access to healthcare ^15^. Considering the above, we conducted a systematic review of literature to explore the roles of CHWs in promoting oral health in SSA and the impact of their interventions.

### Objectives

1. To explore the roles of CHWs in oral health in SSA.
2. To assess the impact of oral health services provided by CHWs.

### Methodology

This systematic review was conducted in accordance with the PRISMA guidelines ^16,17^. Electronic database searches were carried out to identify articles for review. The databases searched included PubMed, Web of Science, Medline, and CINAHL. MeSH (medical subject headings) search terms were used to complete the database searches and retrieved articles were assessed for eligibility using the PICOS (Population, Intervention, Comparator, Outcome, Study design) framework as outlined in Table 1.

**Table 1:** PICOS Framework.

|  |  |
| --- | --- |
| **Population** | CHWs in Sub-Saharan Africa |
| **Intervention** | Oral Health services provided by CHWs |
| **Comparator** | None |
| **Outcome** | Improvement in general physical health<br>Burden or oral health conditions<br>Client satisfaction |
| **Study designs** | All study designs |

The MeSH terms were matched with phrases from the PICO framework in Table 1 and included in the search together with free text terms. MeSH terms are a library of medical headings that are used by the PubMed database to categorise research articles. Free text terms are synonyms, abbreviations and alternative spellings of the terms used in Table 2. This list was compiled by the researchers and is displayed in Table 2.

**Table 2:** Keywords for database search.

| Sub Saharan Africa | Oral Health | Intervention | Community Health Workers | Effect |
| --- | --- | --- | --- | --- |
| Africa<br>Southern Africa<br>East Africa<br>West Africa<br>Sub-Sahara | Oral conditions<br>Oral disease<br>Oral disorders<br>Oral health<br>Oral healthcare<br>Oral pathology<br>Mouth diseases<br>Dental diseases<br>Dental health<br>Mouth health<br>Dental conditions | Initiative<br>Treatment<br>Therapy<br>Programme(s)<br>Outreach<br>Scheme(s)<br>support<br>Health information<br>Health provision<br>Health education<br>Health promotion<br>Screening | Community<br>Health Worker<br>Community Health<br>Extension<br>Worker<br>Lay Health Worker<br>Community Health Assistant<br>Village Health Worker<br>Village Health Team(s)<br>Village Health Volunteer(s)<br>Community Oral Health Workers<br>Community Health Outreach<br>Outreach volunteer<br>Health volunteer<br>Community volunteer<br>Health support volunteer<br>Community helper<br>Rural health volunteer | Result<br>Outcome<br>Impact<br>Efficacy<br>Contribution<br>Acceptability<br>Receptiveness |

#### Updating searches

A database search to update results was carried out before the final analysis stage is completed. The initial database search took place in February 2020 while the final search update and analysis of papers was completed in September 2020. An e-mail alert was set up with PubMed, Web of Science and Medline to ensure that the researchers are notified of new articles that would have shown up in the initial search. A final search was be run to ensure that relevant newly published articles are not missed from the review.

#### Managing references

Database search results were screened by title for inclusion independently by two researchers and stored electronically as an easier way to access articles electronically. The selected articles were exported to the researchers Mendeley account, de-duplicated and stored for further screening.

### Inclusion and exclusion criteria

Studies were assessed against the inclusion criteria namely language, population location, type of publication, year of publication, and country of study. Peer reviewed journal articles that were available in English, conducted in sub-Saharan Africa and published between 2000 – 2019 were included. The inclusion and exclusion criteria are outlined in Table 3.

**Table 3:** Inclusion and exclusion criteria.

| **Inclusion Criteria** | **Exclusion Criteria** |
| --- | --- |
| English language | Non-English language |
| Peer reviewed journal articles and grey literature | Opinion pieces/editorials |
| Published between 2000 – 2019 | Other year of publication other than 2000-2019 |
| Sub-Saharan African country | Country other than Sub-Saharan Africa |

Abstract-only documents were not included in the systematic review as the details published in the full paper may differ from those in the abstract ^18^. There were no ongoing studies that matched all the other inclusion criteria found during the literature search. Previous systematic reviews were excluded from this study to avoid replicating the findings of previous systematic reviews ^19^. Opinion pieces and editorials were also excluded from this systematic review. The study selection process was reported using the Preferred Reporting Items for Systematic reviews and Meta-Analyses (PRISMA) flowchart. PRISMA provides researchers with a transparent manner of reporting systematic reviews ^17^.

### Data extraction

Data extraction was conducted to capture all necessary information from the studies identified from the selection process. The data extraction form was designed to record necessary information that relates to the topic and objectives of the study. This included the general identifying facts about an article such as the name of the researcher carrying out the data extraction, the date of data extraction, a code identifying the article, authorship details, article title, citation, type of publication, country where study was conducted as well as funding and declaration of any conflicts of interest. Other details recorded during the data extraction process related to the characteristics of the study and population including aims and objectives of the study, study design, the inclusion and exclusion criteria, and comparators used. In addition, the form captured the setting of the intervention and any differences between the intervention and control groups.

### Analysis of findings

Subgroup analysis was based on the type of service provided (primary, secondary, tertiary preventive and curative or combination), and by programme aim/goal. The impact of CHWs services was assessed using the core elements of the novel definition for oral health care ^1^ as improved physiological or psychological function or as a reduction in disease or condition status (severity). Disease reduction was measured as decrease in the burden (prevalence, incidence, morbidity/mortality) of any priority oral health condition: tooth decay and cavities (dental caries); gum (periodontal) diseases; oral cancers; oral manifestations of HIV and AIDS; oro-facial trauma from accidents and violence; cleft lip and palate ^20^. An improvement in the oral health related quality of life was also used in measuring programme effectiveness ^21^.

### Ethics

This study involved review of published literature and prior ethical approval for the study was not required.

### Patient and public involvement

There were no patients involved in the conduct of this scoping review.

## Results

### Selection of studies

The PRISMA flow diagram in figure 1 below outlies the process followed during the selection of studies for this review.

**Figure 1:**
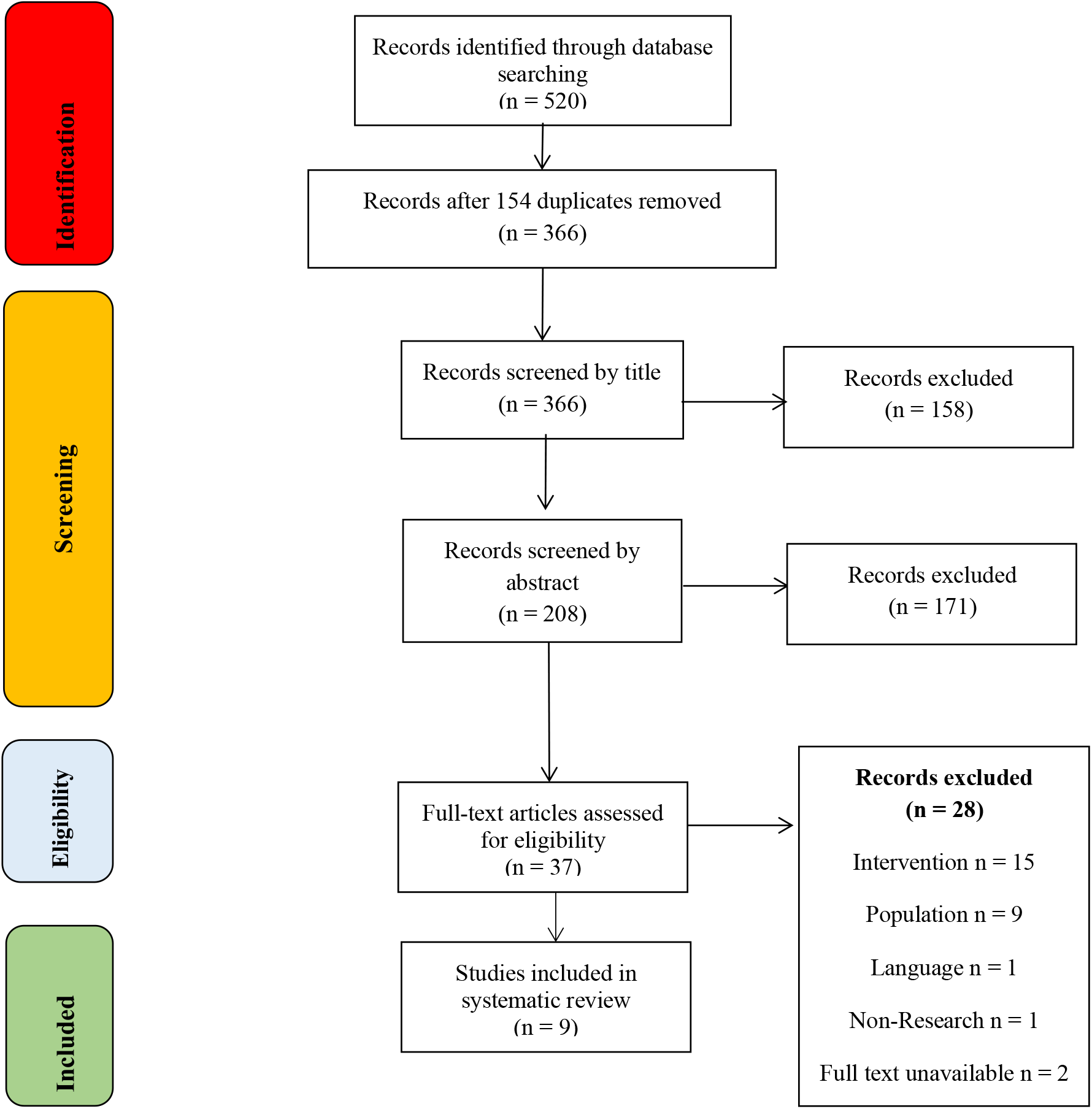
PRISMA flow diagram

This review included nine studies from Kenya ^13^, Cameroon (two studies) ^22,23^, Gambia ^24^, South Africa ^25–27^, Zimbabwe ^28^, and Uganda ^29^. The study designs used across the studies varied and included case control ^13^, cross-sectional ^22,23,25,26^, prospective observational ^24^, experimental ^28,29^, and qualitative ^27^ study designs. Due to the diverse nature of the study designs, targeted population, settings, interventions, and outcomes, a meta-analysis was excluded, and results analysed through narrative synthesis.

### Results synthesis

Various personnel providing oral health services across sub-Saharan African countries met the criteria for inclusion as community health workers. This diverse group of personnel was known by different names such as traditional healers in Cameroon, Uganda, and South Africa ^22,23,25,26,29,30^. Additional titles for these personnel include community health workers in Kenya ^13^, community oral health workers in Gambia ^24^, caregivers ^25^ and oral health promotion officers in South Africa ^27^. There was no standardised pre-service training for CHWs on oral healthcare, hence, their diverse qualifications, training, and scope of their roles. A total population of 2099 CHWs were involved across five studies ^22,23,25,26,29^ while the remainder of the studies did not specify the numbers of CHWs involved in provision of the oral health services ^13,24,27,28,30^.

### Training of CHWs

CHWs training and/ or level of education varied across the included studies with 77.6% of the interventions involving some form of training while 22.2% lacked a training component. The studies ^13,22–25,27,28^ with a training component prior to the CHWs performing their roles aimed at improving the knowledge and skills of CHWs in undertaking oral health interventions whereas the remainder of the studies [7], [10] only entailed observing the behaviour, assessing the knowledge, and practices of CHWs without any form of training for the CHWs. Only four studies specified the training duration for the CHWs prior to their roles and this ranged between a day up to 90 days ^24,25,28,29^. The training for the CHWs was delivered through workshops that entailed focus group discussions, demonstrations including the use of pictorials, and role plays.

The largest (79.87%) improvement in the knowledge of CHWs on common health conditions was reported from a workshop training intervention involving traditional healers in Cameroon ^23^. Improvement in CHWs knowledge on common oral conditions was also reported by a Kenya-based study that trained CHWs on recognizing oral lesions associated with HIV infection ^13^ and a Uganda-based study whereby traditional healers participated in focus group discussions (FGDs) and workshops on false teeth *“ebino”* extractions ^29^ but this improvement was not specified in absolute proportions ^13,29^.

### Roles performed by CHWs in oral health

The findings of this review indicate that CHWs perform a wide range of roles in oral health spanning across the three levels of disease prevention namely primary ^26–29^, secondary ^13,25,26^ and tertiary prevention ^22–24,26,30^ as shown in Figure 2 below and Table 4 above. The roles undertaken by CHWs generally include screening, recognising and offering remedies for common dental problems (e.g., oral candidiasis, dental caries, oral lesions, ulcers, Karposi’s Sarcoma, etc), pain management, atraumatic restorative treatment, teeth extractions, providing health education, basic oral hygiene services, and referring patients for advanced dental services.

**Table 4:** CHWs roles.

| Reference | Study design/ Methods | Country | Population/ setting | No. of CHWs | CHWs role | Service type |
| --- | --- | --- | --- | --- | --- | --- |
| <sup>13</sup> | Case control | Kenya | CHWs | Not specified | Assessing patients.<br>Recognising oral lesions<br>Identifying high-risk patients.<br>Referrals for HIV testing. | Secondary |
| <sup>23</sup> | Cross-sectional | Cameroon | Traditional healers | 21 | Recognising<br>Treating common dental illnesses,<br>Referring patients for advanced dental services. | Tertiary |
| <sup>24</sup> | Prospective Observational | Gambia | Community Oral Health Workers (COHWs) | Not specified | Performing ART under supervision | Tertiary |
| <sup>25</sup> | Cross-sectional | South Africa | Traditional Healers (THs)<br>Caregivers | 32<br>17 | Recognize oral lesions.<br>Oral hygiene (toothbrushing) skills | Secondary<br>Primary |
| <sup>28</sup> | Experimental | Zimbabwe | School teachers | Not specified | Facilitating oral health workshops | Primary |
| <sup>22</sup> | Cross-sectional design | Cameroon | Traditional healers | 16 | Tooth extractions | Tertiary |
| <sup>29</sup> | Pre-, intervention, and follow-up<br>Qualitative | Uganda | Traditional healers<br>Local women | 46<br>1874 | Management of “*ebino*”/ false teeth | Primary |
| <sup>27</sup> | Qualitative | South Africa- | Health promotion officers | 10 | Improving oral health literacy | Primary |
| <sup>26</sup> | Cross-sectional descriptive | South Africa | Traditional healers | 83 | Recognising oral conditions<br>Treatment of common conditions<br>Referral of patients<br>Health education | Primary<br>Secondary<br>Tertiary |

**Figure 2:**
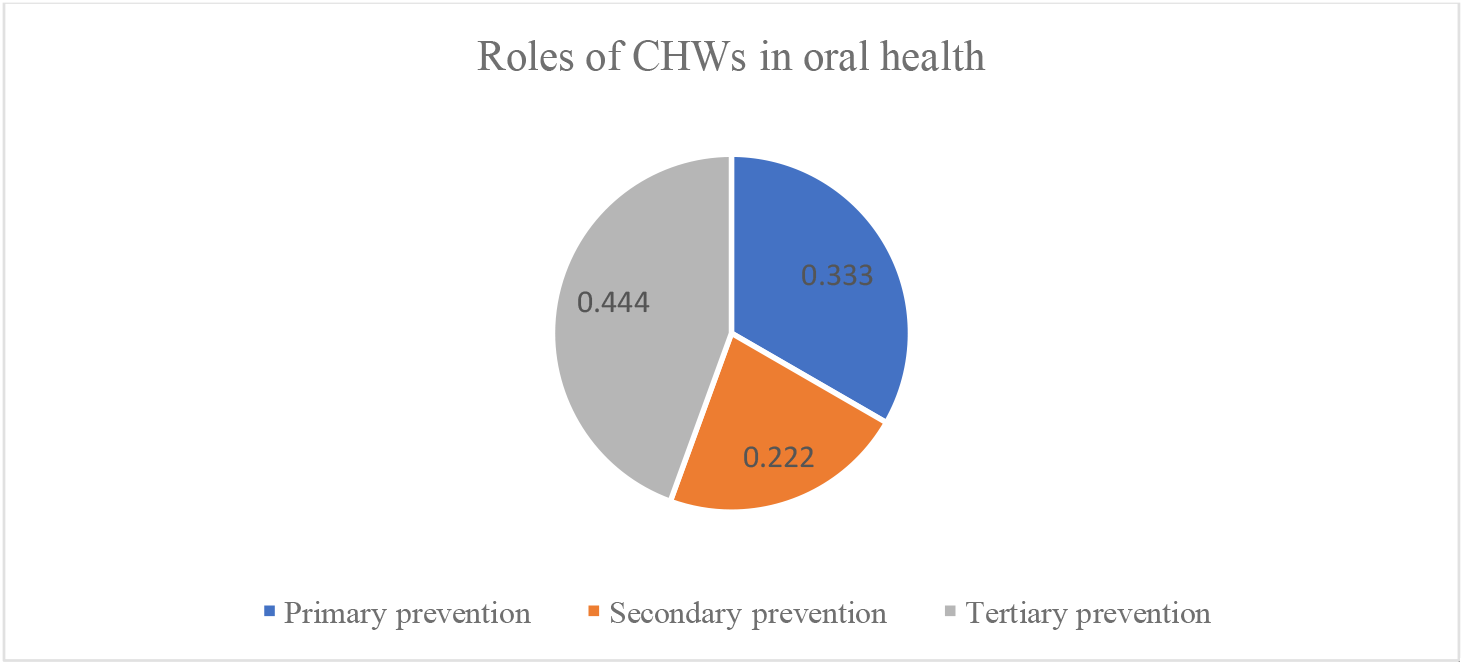
Roles of CHWs in oral health

### Primary prevention

A Zimbabwe-based experimental study involved school teachers who were trained on prevention and treatment of dental caries and periodontal disease, dental fluorosis, and emergency tooth care at school with emphasis on plaque control through the use of the toothbrush and chewing stick in combination with fluoridated toothpaste ^28^. Subsequently, the trained teachers organized regional oral health workshops with 965 school children who were then followed-up in subsequent years to assess for plaque accumulation ^28^. Another study involved traditional healers (46) and local women (1874) in Uganda who participated in focus group discussions and an education session on extraction of false teeth *“ebino”* ^29^. The causes, management, and alternatives to *“ebino”* extractions were also discussed through role plays, dialogues, and pictorials.

The third study involved ten health promotion officers in South Africa who went through a formal training session at Wits University on a revised health promotion curriculum before being subjected to qualitative telephone interviews ^27^. The revised training curriculum was aimed at integrating oral health literacy in the activities of health promoters in the Gauteng region of South Africa ^27^. Another study based in South Africa assessed the knowledge, attitudes and practices of 83 traditional healers on their oral health interventions ^26^. This was a multi-level prevention intervention that entailed primary, secondary, and tertiary prevention activities. The traditional healers provided health education to their patients on toothbrushing, recognised common oral conditions and provided a therapeutic remedies specifically “*muti”* (traditional medicine). Furthermore, the traditional healers also referred the patients for advanced management for their oral conditions ^26^.

### Secondary prevention

A case-control study based in Kenya involved CHWs trained on the recognition of oral lesions associated with HIV infection ^13^. In turn, the trained CHWs would undertake patient assessment within community settings based on the patient’s histories and presenting complaints ^13^. Subsequently, the CHWs identified high-risk patients and referred them appropriately for further testing ^13^. Another study in South Africa also focussed on the oral manifestations of HIV infections where 32 traditional healers and 17 caregivers were part of a focus-group discussion on oral lesions ^25^. The study participants completed a pre- and post-training survey on recognizing oral lesions associated with HIV based on A4-size photographs. In addition, the researchers also assessed the competence of the participants in oral hygiene (toothbrushing skills) ^25^.

### Tertiary prevention

A cross-sectional study assessed the knowledge and practices of 21 traditional healers in north-west Cameroon following a training workshop on recognising, treating, and referring patients with common oral problems ^23^. A follow-up session after three months assessed the knowledge and practices of these traditional healers functioning as CHWs in recognising dental caries, oral candidiasis, aphthous ulcers, Karposi’s sarcoma, and oral cancers as well as the treatments they offered and the associated costs ^23^. Another study based in Cameroon examined the practices of 16 traditional healers regarding tooth extractions in a sample of 150 patients ^22^. The researchers observed the traditional healers as they collected herbs and applied them on the patient’s teeth, extracted teeth, conducted observations post-tooth extraction, and providing post-procedure instructions ^22^. The traditional healers were then asked to complete a questionnaire on the anatomy of the tooth, post extraction instructions, management of complications and prevention of infection ^22^. The researchers also administered questionnaires to patients immediately after undergoing treatment ^22^. In Gambia, community oral health workers (COHWs) were trained for three months on atraumatic restorative treatment (ART) and supervised to provide the treatments ^24^. The ARTs were performed by groups of 10 COHW trainees, and patient outcomes after twelve months compared with ARTs performed by groups of 7 experienced COHWs or 2 dentists ^24^.

### Impact of CHWs interventions

The researchers examined various outcomes to determine the impact of CHWs interventions in oral health. The specific outcomes across the studies included CHWs knowledge on common oral conditions, patients screened for oral ailments, patients referred for advanced treatment, common oral conditions recognised by CHWs, oral health literacy, patients treated by CHWs, dental pain management, patients with complications following dental procedures, preference for CHWs services, patients’ satisfaction with CHWs services, and the burden of dental diseases. Table 5 below summarises the key outcomes as reported across the included studies and these formed the basis for assessing the impact of the roles performed by CHWs in the oral health of the service users.

**Table 5:** Outcomes of CHWs interventions.

| References | 13 | 23 | 24 | 26 | 25 | 28 | 22 | 29 | 27 |
| --- | --- | --- | --- | --- | --- | --- | --- | --- | --- |
| No. of CHWs | Not available | 21 | Not specified | 83 | 49 | Not specified | 16 | 46 | Not specified |
| Intervention | Training | Training | Training | None | Training/FGDs | Training | None | Training/ FGDs | Training |
| Intervention duration (days) | Unavailable | Unavailable | 90 | Unavailable | 2 | 3 | Unavailable | <1 | Unavailable |
| **CHWs roles** |  |  |  |  |  |  |  |  |  |
| Screening for oral diseases | ✓ |  |  |  |  |  |  |  |  |
| Recognising common conditions |  | ✓ |  | ✓ | ✓ |  |  |  |  |
| Treating common conditions | ✓ | ✓ |  | ✓ |  |  |  |  |  |
| Pain management |  |  |  |  |  |  |  |  |  |
| ART |  |  | ✓ |  |  |  |  |  |  |
| Tooth extractions |  |  |  |  |  |  | ✓ | ✓ |  |
| Health education |  |  |  |  | ✓ | ✓ |  |  | ✓ |
| Basic oral hygiene |  |  |  | ✓ | ✓ |  |  |  |  |
| Referral | ✓ | ✓ |  |  |  |  |  | ✓ |  |
| Follow-up (in months) | Not specified | 3 | 12 | Not specified | <1 | 48 | Not specified | 18 | Not specified |
| **Outcomes** |  |  |  |  |  |  |  |  |  |
| CHWs knowledge (%) | ↑ | ↑79.87% | Unavailable | Unavailable | Unavailable | Unavailable | Unavailable | ↑ | Unavailable |
| Patients screened | ↑ | N/A | N/A | Unavailable | Unavailable | N/A | Unavailable | Unavailable | N/A |
| Referred patients | ↑ | Unavailable | N/A | 84% | Unavailable | N/A | 62.5% | ↓ | Unavailable |
| Conditions recognised | Unavailable | Unavailable | N/A | ↑ | ↑ 22.4-71.4% | N/A | Unspecified | Unspecified | N/A |
| Oral health literacy | N/A | N/A | N/A | Unspecified | N/A | ↑ | Unspecified | Unspecified | ↔ |
| No. or % of Patients treated | Unavailable | Unavailable | 131 | Unspecified | N/A | N/A | Unavailable | Unavailable | N/A |
| Pain control effectiveness | N/A | N/A | N/A | Unavailable | N/A | N/A | Unspecified | Unspecified | N/A |
| Patients with complications (%) | Unavailable | Unavailable | Unspecified | Unspecified | N/A | N/A | 4.7% | Unavailable | N/A |
| Preference for CHWs services (%) | Unavailable | 69% | Unavailable | Unspecified | N/A | N/A | 60% | Unspecified | N/A |
| Patient satisfied with CHWs services (%) | Unavailable | 67.3% | Unavailable | Unspecified | N/A | N/A | 93.3% | Unspecified | N/A |
| Burden of oral diseases | Unavailable | Unavailable | Unavailable | Unavailable | Unspecified | ↔ | Unavailable | ↓ | Unspecified |

Legend: ↑: Significant increase; ↓: Significant reduction; ↔: No significant change.

A study on the knowledge, diagnostic, and treatment practices of CHWs in Cameroon reported a 69% preference and 67.3% satisfaction with services offered by traditional healers due to ease of accessibility and low cost compared to formal oral health services ^23^. Similarly, 60% of patients preferred to have their teeth extracted by traditional healers in Cameroon recorded an estimated 93.3% patient satisfaction with the CHWs services and only 4.7% of the patients reported complications afterwards ^22^. A Kenyan-based study reported an improvement in the number of patients screened for oral conditions as well as improved referral of patients for further testing ^13^. Following two days training and focus group discussions, the proportion of oral lesions recognised by CHWs in a South Africa based study involving traditional healers and caregivers increased by 22.4-71.4%. A Uganda-based study on management of false teeth by traditional healers and local women reported a reduction in hospital referrals for children with complications of false teeth extractions ^29^. The reduction in referrals in this study was desirable because the rampart “*ebino*” extractions contributed to a high number of admissions for complications from the procedures undertaken by traditional healers and local women ^29^.

A training intervention on atraumatic restorative therapy (ART) reported no significant differences in ARTs performed by a group of 10 community oral health worker trainees and a comparative group of 7 experienced community oral health workers. There was no significant difference in plaque accumulation amongst school children over four years after participating in an oral health workshop facilitated by school teachers in Zimbabwe compared to the control group ^28^. A knowledge and practices assessment for 83 traditional healers based in South Africa revealed that half of their clients presented with various complaints namely tooth aches, oral candidiasis, and swollen gums ^26^. Subsequently, 90% of them could correctly identify dental caries, gingivitis and oral candidiasis on A4 sized photographs while approximately 82% of the traditional healers referred their patients for advanced treatment in public health facilities ^26^. Following the intervention, the study reported an improvement in the recognition of oral lesions by traditional healers and an 84% improvement in the referral of patients for further specialised treatment ^26^.

## Discussion

### Summary of evidence

This study established that oral health services at community level in sub-Saharan Africa are provided by various groups of personnel namely traditional healers, health promotion officers, caregivers, or simply community healthcare workers. We categorised all the aforementioned personnel as community health workers in accordance with a recent systematic review that defined a CHW as a paraprofessional or lay person who understands the local culture, might have received some form of training and provides basic health services within their community ^31^. Notably, we found no uniformity in the level of training, qualifications, experience, and the roles undertaken by the CHWs based on the included studies. In more than three quarters of the studies, persons undertaking the roles of community healthcare workers in oral health had some form of training leading to their roles. The findings of this review further revealed that the roles undertaken by CHWs in oral health span across the three levels of disease prevention namely primary, secondary, and tertiary prevention.

The primary prevention roles performed by CHWs in oral health include providing oral health education on oral hygiene, toothbrushing skills, organising workshops to promote oral health literacy, managing false teeth in children. The secondary prevention roles performed by community healthcare workers in sub-Saharan Africa include assessing patients’ histories, screening, and recognising oral lesions and referring high-risk patients for further testing and/ or treatment. In tertiary prevention, the reviewed evidence shows that CHWs perform dental extractions, atraumatic restorative therapy under supervision, and providing pain relief for patients with dental problems. The CHWs also play a role in referring patients who require advanced dental interventions to dental specialists in the formal healthcare service. We also observed that CHWs could potentially play a role in promoting oral health literacy through health education which might have an influence on overall health outcomes ^27,28^.

The impact of the CHWs interventions were assessed using the key outcomes of included studies to determine whether there was an improvement in the physiological or psychological functioning of patients or a reduction in the burden and severity of oral conditions. According to the reviewed evidence, CHWs interventions appear to improve access to screening, diagnostic, and treatment services for oral conditions. However, due to the limited number of studies, we could not determine the lasting impact of these improvements on oral health or to what extent they influence the overall burden of oral conditions in the respective study settings. Improvements in the referral of patients for advanced management reported across two studies ranged between 62.5% to 84%. The researchers further observed that training of CHWs could potentially improve their diagnostic skills as reported in one study where CHWs recognised an additional 22.4-71.4% of oral lesions following a two-day training through focus group discussions ^25^.

We also observed a higher (60-69%) preference for CHWs services in comparison to the formal dental services and a resultant 67.3%-93.3% satisfaction for services provided by CHWs ^22,23^. The higher preference for CHWs services appears to be motivated by their closer proximity within the community, the cost, and effectiveness of their services as perceived by the community members in comparison to the formal dental health service providers. The rate of complications reported from CHWs interventions appears to be low (4.7%) but this was only reported in one study where traditional healers performed tooth extractions ^22^. It is not clear based on the reviewed evidence to ascertain the impact of CHWs interventions on the severity of the oral conditions they managed or the physiological functioning of their patients.

### Practice and research implications

The roles played by CHWs with respect to oral health and hygiene are considerably varied between countries and contexts. Their scope may include providing oral hygiene education, recognising common pathologies, and treating oral lesions. In some circumstances trained administration of tooth extractions, dental pain management, and referral for advanced care are functions of CHWs reported across included studies ^22–24,28,29^. Such remit may have a significant impact on oral health and on the burden of oral conditions. Although we could not establish the extent to which these services impacted the general physical health of patients and the overall burden of oral conditions, scientific evidence shows that such services significantly improve people’s overall quality of life ^32,33^. Similarly, a study based in Brazil examined the effectiveness of a CHW program on oral health promotion and reported improvements in oral health knowledge, tooth-brushing practices, self-efficacy in oral hygiene, and utilisation of existing dental services ^34^. Despite this evidence, there is still need for more robust studies to better understand the exact impact of the oral health services provided by CHWs on the physiological functioning of the service users as well as the overall burden of oral health conditions in sub-Saharan Africa. It may also be worth exploring measures that could be applied to standardise the quality of oral health services provided by CHWs in sub-Saharan Africa.

Considering the reviewed evidence, CHWs could provide a readily available pool of workers that can be capacitated to improve access to oral health services in resource limited settings. Our findings suggest that screening for oral conditions, recognition of common dental ailments and providing basic therapeutic remedies for oral problems could potentially benefit from strengthening the services provided by CHWs. We observed an increase in the detection of oral lesions and referrals for advanced management following a training intervention for CHWs ^13,22,25^ but could not ascertain the statistical significance of this increase. This finding is consistent with recent evidence from a cross-sectional study based in India which revealed that trained CHWs were able to consistently recognise oral lesions in comparison with an onsite specialist ^35^. Sub-Saharan African countries have acute shortages for healthcare workers including dental specialists ^36^. In some instances, populations travel long distances to access the limited dental services translating into huge out-of-pocket healthcare costs and wastage of time in search for essential services further exacerbating health inequalities. CHWs have already shown greater promise for bridging the chronic health human resources challenges based on their application across other services including provision of anti-retroviral treatments (ART) for HIV/AIDs, diabetes, and malaria interventions ^37,38^ and could be beneficial in providing oral health services.

The findings of this review show that there is considerable heterogeneity in terms of the role and function and expectation of CHWs with reference to oral healthcare provision. Current research evidence shows that in those contexts where CHWs are most comprehensively trained, they may be equipped with skills to perform essential dental procedures including tooth extractions, and simple pain relief following a period of formal training and supportive supervision ^24^. As such, capacity building the CHWs to provide oral health services could help overcome accessibility, affordability, and acceptability barriers to dental health services in impoverished and marginalised communities. This is further reinforced by a pilot study that demonstrated a higher acceptability for CHWs oral interventions amongst Chinese Americans ^39^. Our findings also revealed a higher preference for and satisfaction with CHWs services owing to their proximity within communities and affordability in comparison to formal health services. In certain communities, CHWs may be the only accessible healthcare provider, hence, an invaluable resource for bridging the disparities in oral health services by providing culturally competent oral health services ^40^.

## Limitations

This review retrieved a small number of studies which were diverse in their study designs and settings. The actual impact of the services offered by CHWs could not be ascertained due to the diverse methods applied across studies and the lack of uniform outcome measures.

## Data Availability

The datasets generated during and/or analysed during the current study are available from the corresponding author on reasonable request through

## Authors’ contributions

All authors discussed the original idea, the methodology used, and contributed to the protocol. MAK served as the subject expert, and all authors (MAK, BOO, ELM, EK, & AWP) participated in the database searches, screening, data extraction, analysis and report writing. BOO and EK served as the first reviewers, ELM and MAK served as the second reviewer, and AWP arbitrated any differences between the two sets of reviewers.

